# irAE-GPT: Leveraging large language models to identify immune-related adverse events in electronic health records and clinical trial datasets

**DOI:** 10.1101/2025.03.05.25323445

**Authors:** Cosmin A. Bejan, Michelle Wang, Sriram Venkateswaran, Ewa A. Bergmann, Laura Hiles, Yaomin Xu, G Scott Chandler, Sam Brondfield, Jordyn Silverstein, Francis Wright, Kimberly de Dios, Daniel Kim, Eric Mukherjee, Matthew S. Krantz, Lydia Yao, Douglas B. Johnson, Elizabeth J. Phillips, Justin M. Balko, Rajat Mohindra, Zoe Quandt

## Abstract

**Background:** Large language models (LLMs) have emerged as transformative technologies, revolutionizing natural language understanding and generation across various domains, including medicine. In this study, we investigated the capabilities, limitations, and generalizability of Generative Pre-trained Transformer (GPT) models in analyzing unstructured patient notes from large healthcare datasets to identify immune-related adverse events (irAEs) associated with the use of immune checkpoint inhibitor (ICI) therapy.

**Methods:** We evaluated the performance of GPT-3.5, GPT-4, and GPT-4o models on manually annotated datasets of patients receiving ICI therapy, sampled from two electronic health record (EHR) systems and seven clinical trials. A zero-shot prompt was designed to exhaustively identify irAEs at the patient level (main analysis) and the note level (secondary analysis). The LLM-based system followed a multi-label classification approach to identify any combination of irAEs associated with individual patients or clinical notes. System evaluation was conducted for each available irAE as well as for broader categories of irAEs classified at the organ level.

**Results:** Our analysis included 442 patients across three institutions. The most common irAEs manually identified in the patient datasets included pneumonitis (N=64), colitis (N=56), rash (N=32), and hepatitis (N=28). Overall, GPT models achieved high sensitivity and specificity but only moderate positive predictive values, reflecting a potential bias towards overpredicting irAE outcomes. GPT-4o achieved the highest F1 and micro-averaged F1 scores for both patient-level and note-level evaluations. Highest performance was observed in the hematological (F1 range=1.0-1.0), gastrointestinal (F1 range=0.81-0.85), and musculoskeletal and rheumatologic (F1 range=0.67-1.0) irAE categories. Error analysis uncovered substantial limitations of GPT models in handling textual causation, where adverse events should not only be accurately identified in clinical text but also causally linked to immune checkpoint inhibitors.

**Conclusion:** The GPT models demonstrated generalizable abilities in identifying irAEs across EHRs and clinical trial reports. Using GPT models to automate adverse event detection in large healthcare datasets will reduce the burden on physicians and healthcare professionals by eliminating the need for manual review. This will strengthen safety monitoring and lead to improved patient care.

## 1. INTRODUCTION

While immune checkpoint inhibitors (ICIs) have greatly enhanced clinical outcomes across multiple cancer types,^1–6^ their use is often accompanied by immune-related adverse events (irAEs), which occur in 74-90% of patients depending on type of ICI therapy.^7^ These events, which can be severe with combination immunotherapies, affect a broad spectrum of organs, including the colon, liver, lungs, heart, nervous system, skin, and endocrine system.^8–15^ Optimizing the management of irAEs in clinical practice, thereby increasing the safety and effectiveness of cancer immunotherapies, is a major focus in precision oncology research. However, current limitations on the timely and accurate identification of irAEs in clinical settings restrict the implementation of such studies at scale. Major challenges stem from the lack of generalizable methods for identifying all types of toxicities and the difficulties of analyzing dynamically evolving real-world healthcare data (especially unstructured data) such as electronic health records (EHRs) and clinical trials datasets.

Previous work by our groups and others has shown that irAEs are poorly captured by structured EHR data like International Classification of Diseases (ICD) codes.^16–19^ Moreover, specific ICD-10 codes for ICI associated irAEs have only become available recently^20^ and it would take time before they are routinely used to abstract irAE data from large healthcare datasets. In contrast, causal relationships between ICI exposures and their irAEs are better encoded in clinical notes, as they allow healthcare providers to document treatment plans, adverse drug reactions, and justifications for discontinuing medications in plain language and with increased detail.^18,19,21^ Still, the automatic identification of irAEs in clinical text poses significant technical challenges since the mention of an irAE in a patient note does not necessarily imply that the patient is currently experiencing the adverse event. For example, an irAE in clinical text may be marked as unlikely or possible, or presented hypothetically as a potential risk during ICI therapy. Further, even when such an event is positively asserted in clinical notes, its underlying cause may not be linked to an ICI exposure.

Recent progress of generative artificial intelligence (AI) technologies, such as Generative Pre-trained Transformer (GPT),^22–24^ underscores the potential of large language models (LLMs) as a transformative approach for identifying irAEs in patient notes. These models have already shown exceptional proficiency in extracting linguistic patterns that convey complex clinical information including clinical phenotypes, assertion statuses, and relationships between medical concepts (e.g., drug–adverse reaction).^25–29^ In this study, we describe the development and evaluation of irAE-GPT, a holistic system created to automatically identify irAEs in clinical text leveraging the state-of-the-art OpenAI’s GPT models, GPT-3.5, GPT-4, and GPT-4o, and two distinct data sources from multiple institutions: clinical trials and real-world EHR data. Clinical trial datasets summarize safety information–such as irAE symptoms, specific irAE diagnoses, and attributes like therapeutic exposure, treatment duration, and comorbidities– in semi-structured reports using standard terminology. In contrast, real-world EHR data capture a comprehensive, longitudinal record of patients’ health trajectories, encompassing diagnoses, procedures, treatments, medications, encounters, laboratory measurements, clinical notes, images, outcomes and other relevant clinical information over time. The objectives of this study were to: (1) assess the capabilities of GPT models for irAE identification in EHR and clinical trial datasets, (2) explore the limitations of GPT models for this task, and (3) evaluate the generalizability and reproducibility of these models across different data sources originating from multiple institutions.

## 2. METHODS

### 2.1 Study design

This retrospective cohort study utilizes data collected from two large EHR systems, Vanderbilt University Medical Center (VUMC) and University of California San Francisco (UCSF), and from Roche-sponsored clinical trials. The study population included patients who received any of the following ICI therapies, either as monotherapy or in combination regimens: atezolizumab, avelumab, durvalumab, ipilimumab, nivolumab, or pembrolizumab. The main analysis was conducted at the patient level, where, for each patient in the study, irAE-GPT reviewed their corresponding notes to identify all possible irAEs resulting from the patient’s exposure to ICI therapy. This analysis also involved the aggregation of irAE predictions by GPT models across all the notes of the same patient. A secondary analysis was carried out on VUMC data to assess irAE-GPT’s performance in extracting irAEs caused by ICIs at the note level. As patients may experience none, one, or several irAEs following ICI exposure, a multi-label classification approach was adopted to ensure LLMs can identify any combination of irAEs associated with each patient or clinical note. System evaluation was conducted for each available irAE as well as for broader categories of irAEs classified at organ level. The irAEs corresponding to each irAE category are listed in **Table S1**. The institutional review boards at VUMC and UCSF approved this study, and all internal processes were followed to enable secondary use of data from Roche sponsored studies for this analysis.

### 2.2 Data sources

At VUMC, the Synthetic Derivative, a de-identified EHR database of 4 million records, was used to sample patients who underwent ICI therapy between 2009 and 2019. Starting from the first day of ICI administration, the health record data associated with each patient was manually reviewed by clinical experts to label all irAEs experienced by the patient or ‘None’ in the event the patient had no adverse reactions caused by ICI therapy. For each patient, clinical notes timestamped between the first date of ICI exposure and up to six months after the last date of ICI administration were selected for LLM analysis. In addition to patient-level annotation of irAEs, the same labels (i.e., a list of irAEs or ‘None’) were used for note-level annotation on individual notes sampled from 20 patients of the above-mentioned dataset.

UCSF Clinical Data Warehouse (CDW), which contains de-identified EHR data captured after 2012 from approximately 6 million unique patients, was used to construct the UCSF cohort. The study team leveraged structured EHR data to identify 617 patients who were admitted to the hospital within 6 months of ICI exposure. Manual chart review narrowed this cohort to 114 subjects who were deemed definitely or probably admitted for hospitalization due to at least one irAE.^30^ The clinical notes selected for this study included the admission, first and last history and physical (H&P) exam notes for that admission, first and last consult notes from various specialty departments for that admission, and discharge summaries for each hospitalization. Ultimately, the final UCSF cohort included a total of 70 patients with 74 hospital encounters, each having at least one of the specified types of clinical notes. Each patient was annotated with specific types of irAEs including the irAEs requiring admission and any other concurrent irAEs. Basic demographics and other baseline characteristics of each patient were extracted from structured EHR data (e.g., age, sex, race, etc.) and manual annotations (e.g., cancer types, cancer stage, etc.)

Clinical trials data came from seven completed Roche-sponsored clinical studies NCT02450331 (n=809), NCT02031458 (n=659), NCT02108652 (n=429), NCT02486718 (n=507), NCT02008227 (n=613), NCT02302807(n=467) and NCT03024996 (n=778) in genitourinary, renal and lung cancer indications (**Figure S1**). irAEs were extracted from the structured database using a broad search of MedDRA preferred terms and this was compared to results from the LLM analysis of free text narratives (written and reviewed by qualified physicians or scientists based on data captured in the study eCRF and additional information provided by the PIs/study sites) to extract irAEs.

### 2.3 irAE-GPT architecture

irAE-GPT is a high-throughput phenotyping system designed to systematically identify irAEs from EHR and clinical trial datasets. It leverages the Azure OpenAI Service, offering access to private, institutionally managed GPT model instances that ensure security and compliance with legal and business agreements. Central to the LLM-based system is a zero-shot prompt crafted to identify irAEs in a clinical note (**Figure 1**). Specifically, for a given patient note, along with prespecified lists of irAEs and ICI treatments, the prompt systematically queries the LLM to determine whether the note describes the patient experiencing any irAEs caused by exposure to any of their ICI treatments. For easier processing, the LLM output was required to be in JavaScript Object Notation (JSON) format, with each irAE corresponding to a binary response (‘Yes’ or ‘No’) based on its presence in the clinical note. Further, sets of text expressions describing synonyms, acronyms, or semantically related medical concepts–which we called “synsets”–were constructed for selected irAEs to better guide the LLM in the irAE identification process. For example, using the synset information corresponding to ‘*neuropathy’* listed in **Table S2**, its augmented query becomes “*Output ‘Yes’ if the patient has experienced neuropathy (neurotox, neurotoxicity) because of exposure to one or more immune checkpoint inhibitors. Otherwise, output ‘No’.*” Notably, the irAE list was adapted for each institution such that each element from the list was annotated at least once in the corresponding dataset. Similarly, multiple site-specific irAE synsets emerged due to variations in the manual annotation processes at each institution. For example, the prompt for the VUMC run had separate queries for colitis and diarrhea whereas the UCSF prompt had only one query for both concepts because they were annotated under the same irAE label. In this case, the colitis synset for the UCSF prompt contained both ‘*colitis*’ and ‘*diarrhea*’. The python code with the implementation of irAE-GPT is available at https://github.com/bejanlab/irAE-GPT.git.

**Figure 1.**
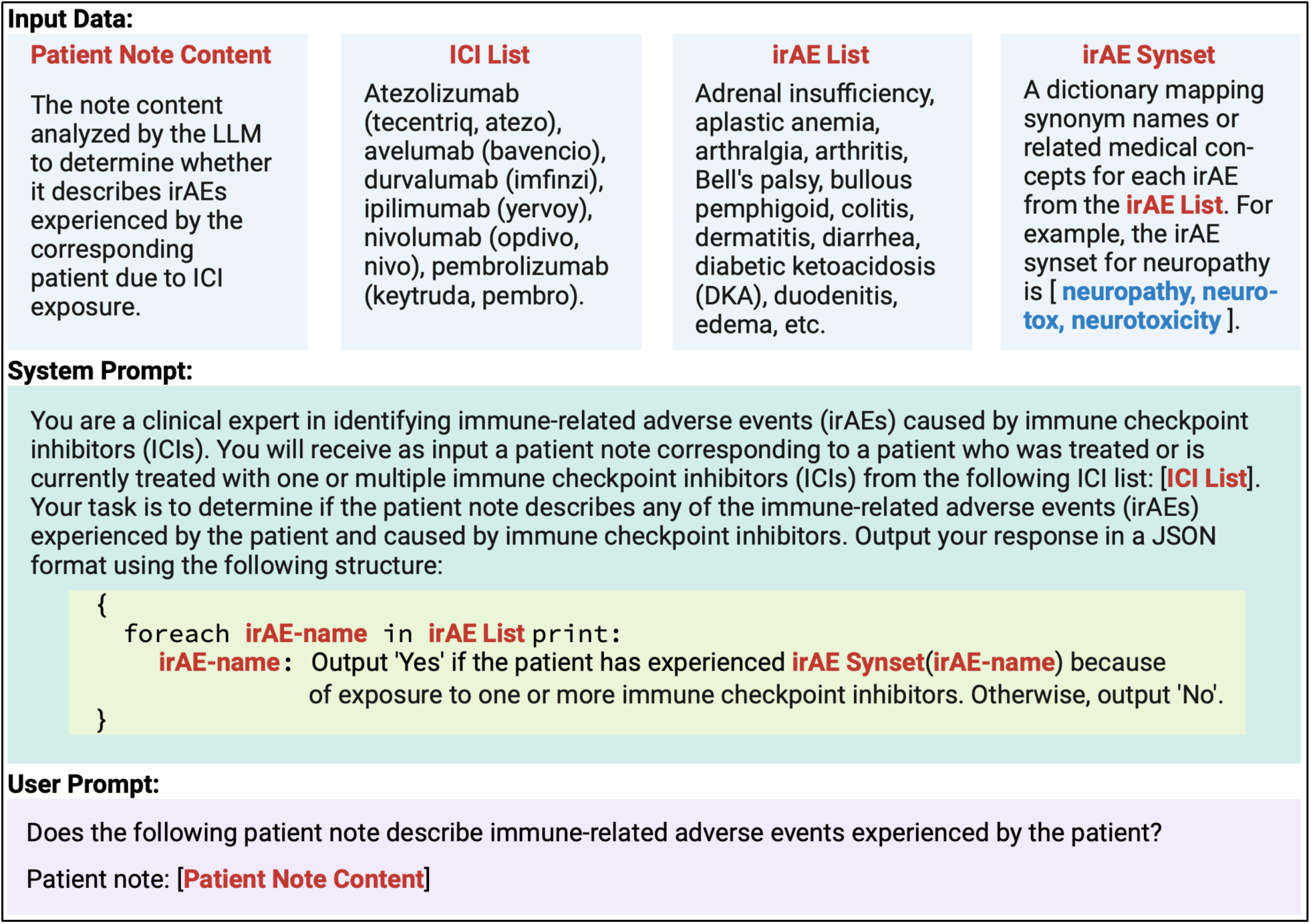
Prompt template for identifying irAEs due to ICI exposure in patient notes. The irAEs used for each dataset are listed in **Tables S3-S5**.

### 2.4 irAE-GPT evaluation

The system was deployed at each institution, and its evaluation utilized the complete dataset from the corresponding institution. The same zero-shot prompt and hyperparameters were used for each run except for site-specific irAE lists and synsets. Each LLM was executed with the temperature–a hyperparameter that controls the level of randomness or determinism in LLM output generation–set to 0. This setting is optimal for binary classification tasks, ensuring the model consistently selects the highest-probability token, corresponding to ‘*Yes*’ or ‘*No*’ for each irAE. Performance metrics including precision (positive predictive value or PPV), recall (sensitivity), specificity, and F1 score were computed by comparing the manual annotations against the results extracted by irAE-GPT for each irAE. Patient-level evaluation involved aggregating the LLM results from all notes per patient and comparing the aggregated results with the annotated irAE labels of the corresponding patient. To extract performance measures for irAE categories, we converted both irAE annotations and irAE results generated by the system using the mappings listed in **Table S1**. Micro- and macro-averaged results were also computed to assess the overall performance across all irAEs and irAE categories. Because our task emphasized the accurate identification of positive outcomes, F1 and micro-averaged F1 were selected as the primary metrics throughout the experiments. Finally, error analysis was conducted based on manual assessment of clinical notes corresponding to false positive and false negative results.

## 3. RESULTS

### 3.1 Description of datasets

The cohorts at VUMC, UCSF, and Roche included 100, 70, and 272 patients, respectively (**Table 1**). The mean age of the patients at ICI initiation across the three cohorts ranged from 62.8 to 65.2 years, with most being male and White. The VUMC and UCSF patients were more likely to be treated with a PD-(L)1 inhibitor followed by a combination of PD-(L)1 and CTLA-4 inhibitors while all Roche patients were treated with a PD-L1 inhibitor. Melanoma was the most common cancer type among VUMC and UCSF patients, while lung cancer was the most frequent in the selected Roche-sponsored clinical trials. Each institute adopted its own strategy in annotating irAEs and selecting unstructured patient notes for LLM-based analysis. At VUMC, the 100-patient cohort was selected from a total of 747 patients who were exposed to ICI therapy and annotated with irAEs (**Table S3**). Since there were no restrictions imposed on the ICI exposed patients for irAE annotation, many patients were found as not experiencing any irAE (42 patients labeled as ‘None’ out of 100 patients selected for the study). From the patients with at least one irAE, most of them had rash (N=10), colitis (N=9), and pneumonitis (N=8). As no restriction was imposed on patient note selection either (e.g., by note type), 26,432 of them (264 notes per patient, on average) were included for LLM-based irAE identification. At UCSF, all 70 patients were found as experiencing at least one irAE as they were selected for annotation from patients admitted for hospitalization due to definite or probable irAE (hence, with more severe events). Gastrointestinal irAEs, mostly colitis (N=23) and hepatitis (N=15), and pneumonitis (N=15) were the most common irAEs at hospitalization (**Tables 1** and **S4**). For LLM-based irAE identification, the study team curated a set of 487 clinical notes from 74 hospital encounters (∼7 notes per patient) corresponding to the 70 patients. Finally, at Roche, there were 3,471 patients who received ICI therapy in 7 completed trials. From the dataset, 272 patients, who corresponded to the same number of reports, were extracted using a broad structured MedDRA search for possible irAEs. A subset of the most common irAEs were selected for the LLM-based analysis (**Figure S1**, **Table S5**). Notably, some adverse event terms extracted during this process may not qualify as irAEs (e.g., influenza) but might instead be indirectly associated with ICI therapy. For instance, influenza-like symptoms may be reported as influenza. In the Roche study cohort, the most common irAEs were pneumonitis (N=41), colitis (N=24), and rash (N=15).

**Table 1.**
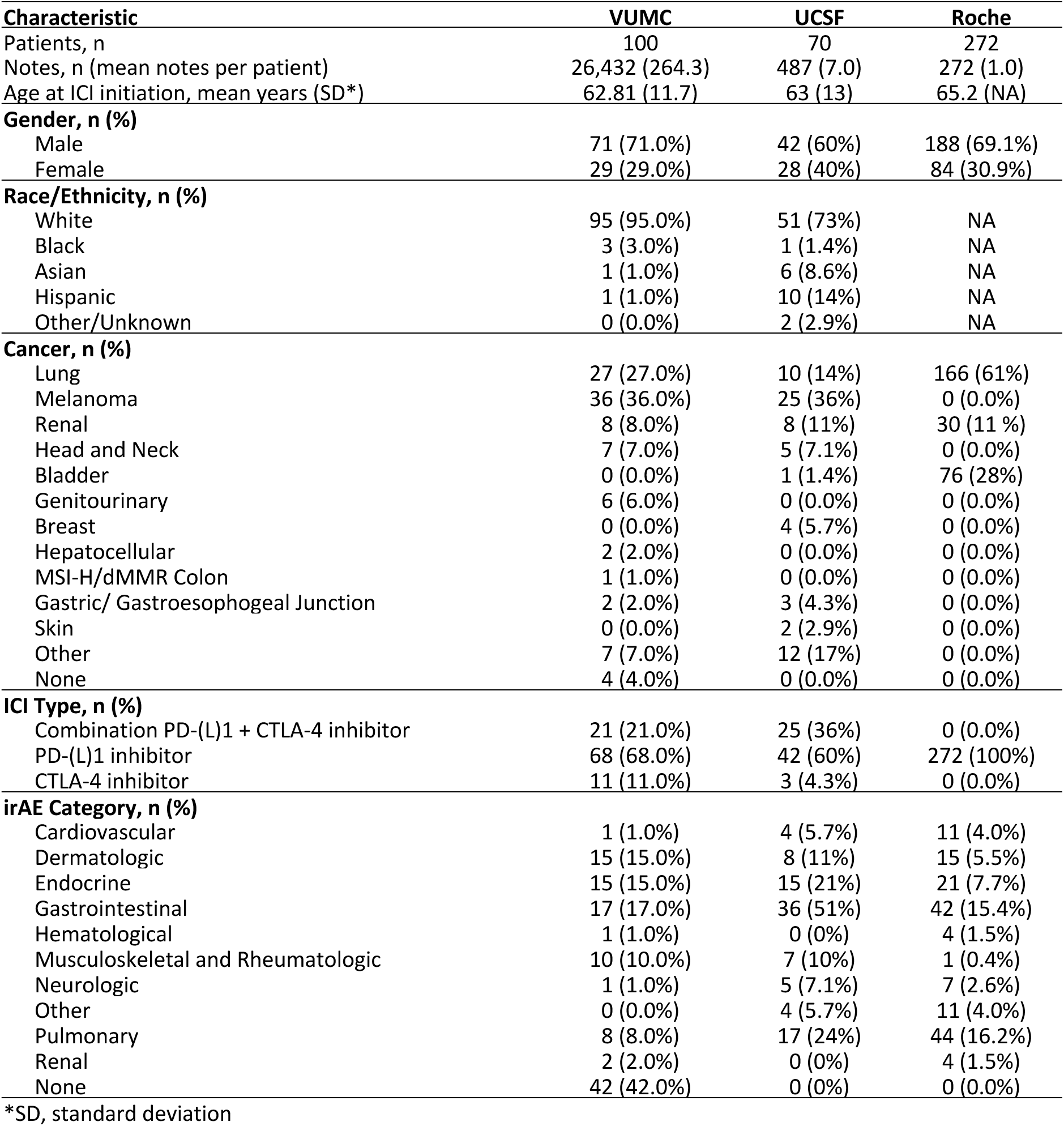
Selected characteristics of study participants across all sites.

### 3.2 Patient-level evaluation

We adopted simple strategies to aggregate the predictions for all the notes of the same patient. For VUMC results, we balanced the tradeoff between precision and recall by determining the optimal decision threshold—the number of notes with irAE-positive predictions—used to classify patient-level irAE outcomes. The optimal decision threshold values determined by GPT-3.5, GPT-4, and GPT-4o models for maximizing the micro-averaged F1 score were 6, 10, and 18, respectively (**Figures S2-S4**). The patient-level irAE predictions on UCSF and Roche datasets were determined based on at least one positive prediction (decision threshold=1) in any of the notes corresponding to the same patient.

Overall, the GPT models exhibited high recall (sensitivity) and specificity but achieved only moderate precision (positive predictive value, PPV) across datasets from all institutions (**Tables S6-S8**). For multiple irAEs at VUMC (e.g., hemolytic anemia, lupus flare, bullous pemphigoid), UCSF (e.g., Stevens-Johnson syndrome, Guillain-Barre), and Roche (e.g., bladder tamponade, ascending flaccid paralysis), the GPT models achieved 100% F1 scores. However, the models’ performance was relatively modest for several more common adverse events including arthritis (*F*1*_VUMC_*_:*GPT*-4_ = 0.22, *F*1*_UCSF_*_:*GPT*-3.5_ = 0.27), rash (*F*1*_VUMC_*_:*GPT*-3.5_ = 0.31), colitis (*F*1*_VUMC_*_:*GPT*-4*o*_ = 0.32), fever (*F*1*_UCSF_*_:*GPT*-4*o*_ = 0.32), hypothyroidism (*F*1*_Roche_*_:*GPT*-4_ = 0.31), and pyrexia (*F*1*_Roche_*_:*GPT*-4*o*_ = 0.33). The overall trends in primary measures, F1 and micro-averaged F1, indicate GPT-4o as the best performing model.

Organ-level classification of irAEs facilitates the comparison of results both across institutions and within individual irAE categories (**Figure 2** and **Table 2**). Across the VUMC, UCSF, and Roche datasets, the best results were observed in the hematological (F1 range=1.0-1.0), gastrointestinal (F1 range=0.81-0.85), and musculoskeletal/rheumatologic (F1 range=0.67-1.0) categories, respectively. GPT models showed consistently high F1 scores in the pulmonary category across all datasets. Notably, aggregating manually annotated irAEs and irAE predictions into organ-level irAE categories improves the micro-averaged F1 scores across all datasets (**Tables S6-S8**).

**Figure 2.**
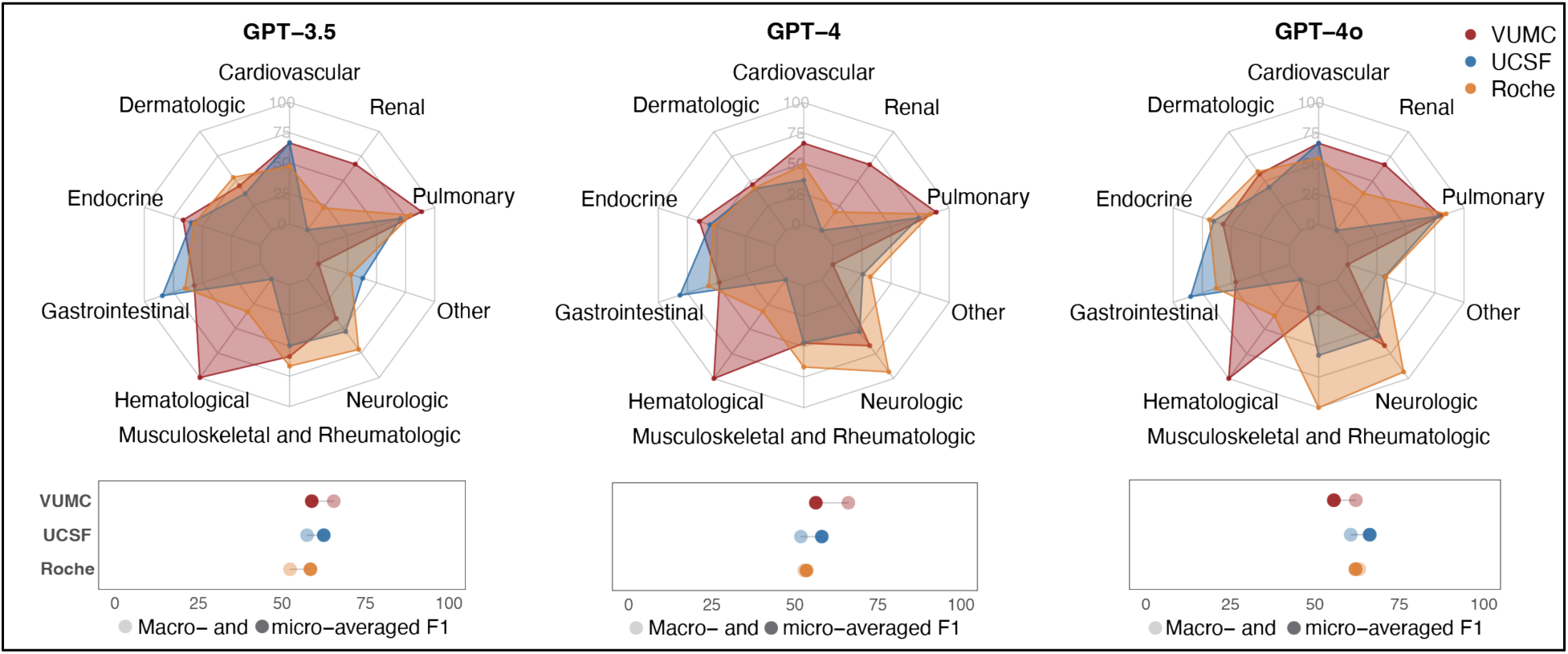
Comparative analysis of patient-level results across datasets from 3 institutions: Vanderbilt University Medical Center (VUMC), University of California San Francisco (UCSF), and Roche. The radar charts at the top highlight variations in F1 scores across irAE categories, whereas the lollipop plots at the bottom summarize the macro- and micro-averaged F1 scores. Detailed results of this analysis are provided in **Table 2**.

**Table 2.**
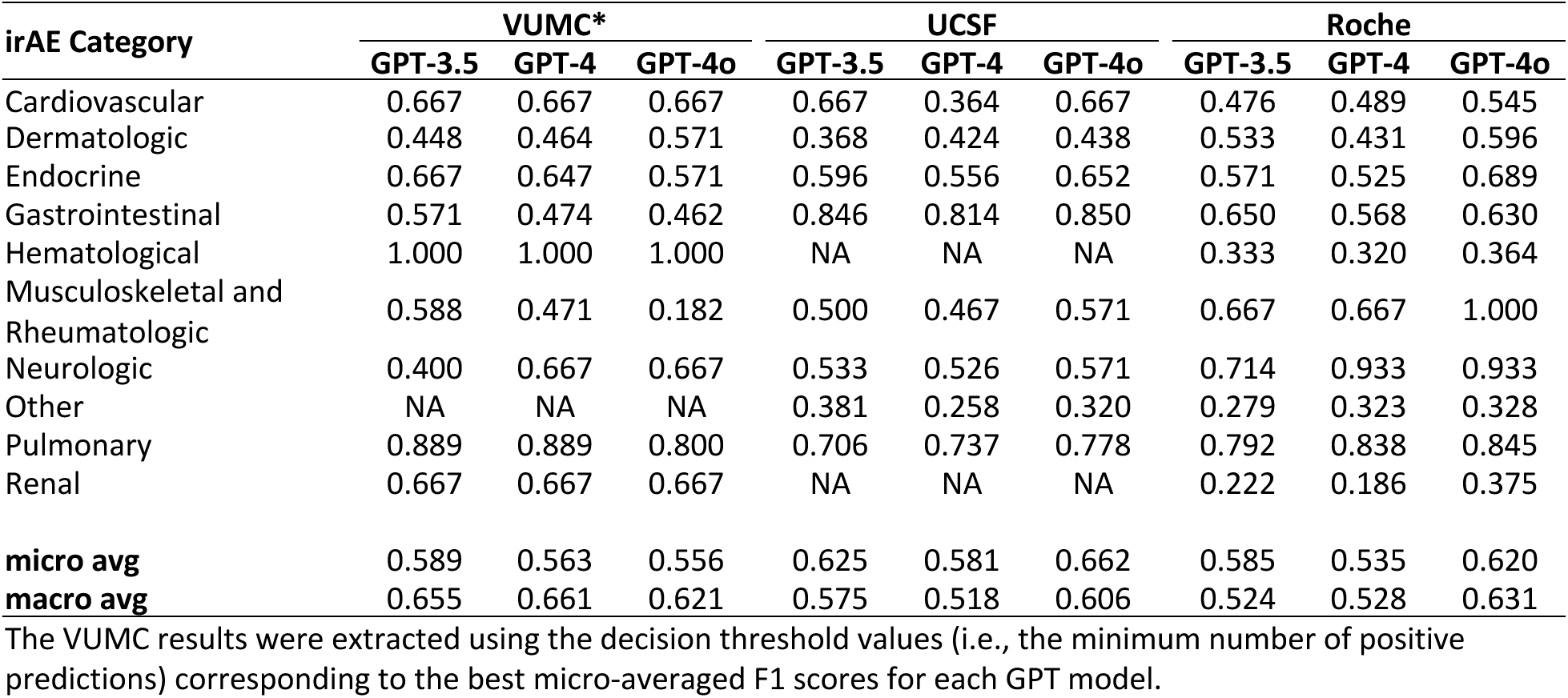
Patient-level evaluation for the identification of irAE categories across the datasets from Vanderbilt University Medical Center (VUMC), University of California San Francisco (UCSF), and Roche. All the result values in the table correspond to F1 scores.

| irAE Category | VUMC* |  |  | UCSF |  |  | Roche |  |  |
| --- | --- | --- | --- | --- | --- | --- | --- | --- | --- |
|  | GPT-3.5 | GPT-4 | GPT-4o | GPT-3.5 | GPT-4 | GPT-4o | GPT-3.5 | GPT-4 | GPT-4o |
| Cardiovascular | 0.667 | 0.667 | 0.667 | 0.667 | 0.364 | 0.667 | 0.476 | 0.489 | 0.545 |
| Dermatologic | 0.448 | 0.464 | 0.571 | 0.368 | 0.424 | 0.438 | 0.533 | 0.431 | 0.596 |
| Endocrine | 0.667 | 0.647 | 0.571 | 0.596 | 0.556 | 0.652 | 0.571 | 0.525 | 0.689 |
| Gastrointestinal | 0.571 | 0.474 | 0.462 | 0.846 | 0.814 | 0.850 | 0.650 | 0.568 | 0.630 |
| Hematological | 1.000 | 1.000 | 1.000 | NA | NA | NA | 0.333 | 0.320 | 0.364 |
| Musculoskeletal and Rheumatologic | 0.588 | 0.471 | 0.182 | 0.500 | 0.467 | 0.571 | 0.667 | 0.667 | 1.000 |
| Neurologic | 0.400 | 0.667 | 0.667 | 0.533 | 0.526 | 0.571 | 0.714 | 0.933 | 0.933 |
| Other | NA | NA | NA | 0.381 | 0.258 | 0.320 | 0.279 | 0.323 | 0.328 |
| Pulmonary | 0.889 | 0.889 | 0.800 | 0.706 | 0.737 | 0.778 | 0.792 | 0.838 | 0.845 |
| Renal | 0.667 | 0.667 | 0.667 | NA | NA | NA | 0.222 | 0.186 | 0.375 |
| **micro avg** | 0.589 | 0.563 | 0.556 | 0.625 | 0.581 | 0.662 | 0.585 | 0.535 | 0.620 |
| **macro avg** | 0.655 | 0.661 | 0.621 | 0.575 | 0.518 | 0.606 | 0.524 | 0.528 | 0.631 |

The VUMC results were extracted using the decision threshold values (i.e., the minimum number of positive predictions) corresponding to the best micro-averaged F1 scores for each GPT model.

### 3.3 Note-level evaluation

A note-level evaluation was conducted to more effectively assess the robustness of GPT models, given that patient-level outcomes depend on the aggregation of irAE predictions across all notes for the same patient. Further, this type of approach facilitates the identification of irAEs over time, offering direct utility for incident-based pharmacoepidemiologic studies. Overall, despite finding optimal decision thresholds for patient-level predictions, higher micro-averaged F1 scores were observed in note-level evaluation compared to patient-level evaluation for identifying both irAEs (micro-averaged F1 ranges, [0.50-0.57] vs. [0.46-0.48]) and irAE categories (micro-averaged F1 ranges, [0.61-0.66] vs. [0.56-0.59]) in the VUMC dataset (**Tables 3** and **S6**). Consistent with the trends shown for patient-level evaluation, GPT-4o yielded the best performing F1 scores for note-level evaluation.

**Table 3.** Note-level evaluation for the identification of irAEs and their corresponding categories in the Vanderbilt University Medical Center (VUMC) dataset.

| irAE | GPT-3.5 |  |  |  | GPT-4 |  |  |  | GPT-4o |  |  |  |
| --- | --- | --- | --- | --- | --- | --- | --- | --- | --- | --- | --- | --- |
|  | P | R | S | F1 | P | R | S | F1 | P | R | S | F1 |
| Neuropathy | 0.217 | 0.098 | 0.971 | 0.135 | 0.553 | 0.412 | 0.972 | 0.472 | 0.600 | 0.294 | 0.984 | 0.395 |
| Hypothyroid | 0.431 | 0.936 | 0.906 | 0.591 | 0.419 | 0.936 | 0.902 | 0.579 | 0.688 | 0.936 | 0.968 | 0.793 |
| Myasthenia gravis (MG) | 0.902 | 0.979 | 0.992 | 0.939 | 0.885 | 0.979 | 0.990 | 0.929 | 0.979 | 0.979 | 0.998 | 0.979 |
| Rash | 0.535 | 0.676 | 0.968 | 0.597 | 0.351 | 0.794 | 0.921 | 0.486 | 0.386 | 0.794 | 0.932 | 0.519 |
| Colitis | 0.714 | 0.741 | 0.988 | 0.727 | 0.724 | 0.778 | 0.988 | 0.750 | 0.750 | 0.778 | 0.989 | 0.764 |
| Adrenal insufficiency | 0.451 | 0.958 | 0.956 | 0.613 | 0.431 | 0.917 | 0.955 | 0.587 | 0.431 | 0.917 | 0.955 | 0.587 |
| Hepatitis | 0.750 | 0.750 | 0.991 | 0.750 | 0.655 | 0.792 | 0.984 | 0.717 | 0.633 | 0.792 | 0.983 | 0.704 |
| Arthralgia | 0.208 | 1.000 | 0.870 | 0.344 | 0.321 | 0.818 | 0.941 | 0.462 | 0.396 | 0.955 | 0.950 | 0.560 |
| Duodenitis | 0.778 | 0.933 | 0.994 | 0.848 | 0.833 | 1.000 | 0.995 | 0.909 | 0.833 | 1.000 | 0.995 | 0.909 |
| Pancreatitis | 0.706 | 0.923 | 0.992 | 0.800 | 0.722 | 1.000 | 0.992 | 0.839 | 0.722 | 1.000 | 0.992 | 0.839 |
| Hypophysitis | 0.279 | 1.000 | 0.953 | 0.436 | 0.234 | 0.917 | 0.945 | 0.373 | 0.239 | 0.917 | 0.947 | 0.379 |
| Mucositis | 1.000 | 0.583 | 1.000 | 0.737 | 0.526 | 0.833 | 0.986 | 0.645 | 0.818 | 0.750 | 0.997 | 0.783 |
| Arthritis | 0.200 | 0.778 | 0.957 | 0.318 | 0.179 | 0.778 | 0.951 | 0.292 | 0.286 | 0.667 | 0.977 | 0.400 |
| Pneumonitis | 0.350 | 0.875 | 0.980 | 0.500 | 0.318 | 0.875 | 0.977 | 0.467 | 0.316 | 0.750 | 0.980 | 0.444 |
| Joint pain | 0.037 | 1.000 | 0.883 | 0.071 | 0.067 | 1.000 | 0.937 | 0.125 | 0.045 | 1.000 | 0.904 | 0.086 |
| Fever | 0.111 | 0.667 | 0.976 | 0.190 | 0.071 | 0.667 | 0.961 | 0.129 | 0.080 | 0.667 | 0.965 | 0.143 |
| Myalgia | 0.036 | 0.667 | 0.920 | 0.069 | 0.057 | 0.667 | 0.950 | 0.105 | 0.049 | 0.667 | 0.941 | 0.091 |
| **micro avg** | 0.370 | 0.754 | 0.959 | 0.496 | 0.407 | 0.814 | 0.962 | 0.542 | 0.445 | 0.797 | 0.968 | 0.571 |
| **macro avg** | 0.453 | 0.798 | 0.959 | 0.510 | 0.432 | 0.833 | 0.962 | 0.521 | 0.485 | 0.815 | 0.968 | 0.551 |
| **irAE Category** |  |  |  |  |  |  |  |  |  |  |  |  |
| Dermatologic | 0.535 | 0.676 | 0.968 | 0.597 | 0.351 | 0.794 | 0.921 | 0.486 | 0.386 | 0.794 | 0.932 | 0.519 |
| Endocrine | 0.580 | 0.945 | 0.916 | 0.719 | 0.535 | 0.945 | 0.899 | 0.683 | 0.793 | 0.945 | 0.970 | 0.862 |
| Gastrointestinal | 0.831 | 0.771 | 0.982 | 0.800 | 0.714 | 0.857 | 0.960 | 0.779 | 0.766 | 0.843 | 0.970 | 0.803 |
| Musculoskeletal and Rheumatologic | 0.280 | 0.971 | 0.866 | 0.434 | 0.264 | 0.971 | 0.855 | 0.415 | 0.288 | 0.941 | 0.875 | 0.441 |
| Neurologic | 0.859 | 0.561 | 0.984 | 0.679 | 0.791 | 0.694 | 0.968 | 0.739 | 0.847 | 0.622 | 0.981 | 0.718 |
| Other | 0.111 | 0.667 | 0.976 | 0.190 | 0.071 | 0.667 | 0.961 | 0.129 | 0.080 | 0.667 | 0.965 | 0.143 |
| Pulmonary | 0.350 | 0.875 | 0.980 | 0.500 | 0.318 | 0.875 | 0.977 | 0.467 | 0.316 | 0.750 | 0.980 | 0.444 |
| **micro avg** | 0.544 | 0.759 | 0.953 | 0.634 | 0.483 | 0.831 | 0.934 | 0.611 | 0.555 | 0.800 | 0.953 | 0.656 |
| **macro avg** | 0.507 | 0.781 | 0.953 | 0.560 | 0.435 | 0.829 | 0.934 | 0.528 | 0.497 | 0.795 | 0.953 | 0.562 |

P, precision (positive predictive value); R, recall (sensitivity); S, specificity; F1, F1-measure.

### 3.4 Error analysis

The manual review of notes associated with false positive results revealed that LLMs struggle to solve causal relationships involving positively asserted events with unspecified or unrelated etiological factors. False positive findings also included adverse events characterized by an assertion status indicating hypothetical, uncertain or possible scenarios, or events not associated with the patient. **Table S9** lists examples with clinical note excerpts where such adverse events were misclassified by LLMs as irAEs. Finally, several false positives were ultimately confirmed as true irAEs, having been unnoticed during manual annotation (e.g., “*treated with ipilimumab c/b hypophysitis and colitis*”). A significant lower number of false negative results were generated by the GPT models, many of which being improperly described in the zero-shot prompt. For instance, the LLMs failed to associate terms such as “*liver inflammation*” and “*developed grade 2 elevated LFTs*” with hepatitis, as the medical concepts encoded in these expressions were missing from the hepatitis synset. Improper tokenization of specific medical terms may also impact how accurately GPT models recognize irAEs in clinical notes. Subword tokenization examples generated by Tiktoken,^31^ the tokenizer developed by OpenAI for its GPT models, include [*‘neuropathy’*] → [*’neu’, ‘rop’, ‘athy’*], [*‘neurotox’*] → [*’ne’, ‘uro’, ‘to’, ‘x’*], [‘*myasthenia gravis*’] → [*’my’, ‘ast’, ‘hen’, ‘ia’, ‘ gr’, ‘avis’*], and [‘*hypothyroid’*] → [*’h’, ‘yp’, ‘othy’, ‘roid’*].

## 4. DISCUSSION

This study demonstrates that large language models have potential to effectively identify immune-related adverse events (irAEs) in two distinct textual modalities: clinical notes from two large, tertiary, real-world EHR systems and patient reports from seven clinical trials. Moreover, our findings suggest that GPT models are well-suited for exhaustive identification of irAEs across diverse clinical contexts. At VUMC, we evaluated the LLMs in both inpatient and outpatient settings for patients undergoing ICI therapy, regardless of whether they experienced irAEs. The UCSF experiments utilized EHR data from patients who were hospitalized within six months after ICI exposure due to one or more irAEs, which are considered more severe. The experimental setup at Roche relied on data from patients participating in clinical trials on ICI therapy and experiencing at least one irAE. The primary outcome of our study is to demonstrate irAE-GPT as a generalizable, high-throughput system for automatically extracting therapy-associated toxicities from clinical text. The system aims to alleviate the burden of manually annotating irAEs in clinical text, a labor-intensive task requiring experienced and specialized clinicians. It extracts irAE outcomes at both patient and note levels, enabling a wide range of pharmacoepidemiologic studies, from those focusing on prevalence to those addressing incidence, ultimately contributing to guide clinical care to improve safe and optimized use of novel immunotherapies.

The evaluation of the GPT models revealed several significant findings. First, the models achieved consistently high recall/sensitivity and specificity values at the cost of moderate precision scores. This resulted in significantly more false positives than false negatives, indicating a bias in the GPT models toward sensitivity or overpredicting positive outcomes. One of our experiments to mitigate this bias focused on adjusting the decision threshold to optimize the F1 score, achieving a more balanced trade-off between precision and recall. Second, differences in performance were noted among the various types of evaluations conducted. The note-level evaluation yielded better results than the patient-level evaluation, possibly influenced by how note-level predictions were combined to derive patient-level outcomes. Moreover, the prediction of irAE categories demonstrated higher performance values compared to specific irAE evaluation results, primarily due to the relaxed approach in positively predicting the organ-level adverse events. For instance, positively predicting any one of pneumonitis, wheezing, influenza, ARDS, or pleuritis is sufficient to classify the pulmonary category as positive. Finally, the error analysis highlights the challenges faced by LLMs in handling textual causation, a task more complex than named entity recognition, where medical conditions identified in clinical text (e.g., adverse events) must also be linked to specific etiological factors (e.g., immune checkpoint inhibitors).

To the best of our knowledge, this is the first study leveraging OpenAI’s GPT models to identify irAEs in clinical text. The closest study to ours is by Sun et al,^19^ where they employed the open-source LLM Mistral OpenOrca^32^ to detect four irAEs (colitis, hepatitis, myocarditis, and pneumonitis) in electronic health records of hospitalized patients at Massachusetts General Hospital (MGH) and the Brigham and Women’s Hospital (BWH). A straightforward analysis using averaged F1 scores over the four irAEs revealed that Mistral OpenOrca underperformed on the MGH (mean F1=0.26) and BWH (mean F1=0.29) datasets compared to GPT models, which performed better on the VUMC (mean F1=0.60), UCSF (mean F1=0.73), and Roche (mean F1=0.65) datasets. Here, the Roche results reflect the averaged F1 scores for identifying colitis, hepatitis, and pneumonitis, as myocarditis events were absent in the corresponding dataset.

With the anticipated release of new ICD-10 codes for ICI-associated irAEs,^20^ the classification of irAEs is expected to become more precise, leading to improvements in data availability, comparability, and quality. However, the consistent and routine adoption of these new codes by clinicians, as well as their validation in clinical applications, will require time. The synergy between irAE-GPT and the new ICD codes could enable a systematic approach to understanding the spectrum and burden of ICI-associated irAEs, with far-reaching implications for clinical practice, public health, and research.

Our study has several limitations that warrant further investigation in future research work. First, we did not investigate widely-used methods for enhancing the performance of LLMs, including prompt engineering, few-shot learning, and fine-tuning.^23,33,34^ For example, our groups and others have previously demonstrated that LLMs are highly sensitive to the way prompts are constructed, and that prompt augmentation and optimization play a key role in improving the performance of various tasks.^26,33,35,36^ Second, due to the input token limit of GPT models, for the patient level approach, we implemented a strategy to aggregate irAE predictions from all notes belonging to the same patient. This is because, although some GPT models offer relatively large token limits, using prompts that could potentially contain all the notes of a patient is still infeasible. For example, on the VUMC dataset, a patient has on average 264 notes during the ICI exposure period. In future work, we intend to use retrieval-augmented generation (RAG) techniques^37^ as an alternative approach, creating one prompt per patient that contains only irAE-relevant text snippets extracted from the entire patient record. Third, the possibility that LLMs may perpetuate and exacerbate health disparities in clinical studies of cancer immunotherapy safety and effectiveness has not yet been explored. While not within the scope of our study, the implementation of fairness assessments and bias mitigation techniques is critical for ensuring transparent and equitable LLM approaches in such studies. Lastly, the lack of a standardized and comprehensive list of irAEs, combined with challenges in categorizing them through splitting or grouping, may limit the applicability of our findings to other institutions or treatment centers.

## 5. CONCLUSION

This study introduces irAE-GPT, a high-throughput phenotyping system that leverages GPT models to systematically identify irAEs in unstructured patient notes from multiple EHR systems and clinical trials. The GPT models demonstrated robust performance and generalizable capabilities in identifying irAEs across diverse clinical contexts, reducing the need for manual annotation of adverse events in clinical text and highlighting their potential to improve the characterization of the safety profile of cancer immunotherapies. Efforts should continue to enhance the capabilities of these models in identifying textual causation between medication exposures and adverse events, while also addressing their potential biases in extraction of safety data from clinical text. irAE-GPT is publicly available, allowing researchers to experiment with their local healthcare datasets.

## Data availability

The source code used in the analysis is publicly available on GitHub at https://github.com/bejanlab/irAE-GPT.git. The summary statistics extracted for this study are provided in the manuscript and supplementary material. Any request to access the Synthetic Derivative data will need to be reviewed and approved by Vanderbilt University Medical Center. Researchers will need to provide evidence of IRB approval for their study. For the approved studies, data will be released via a Data Use Agreement.

## Funding

CAB, DBJ, and JB are supported by R01CA227481. CAB and JB are supported by R01HL156021. CAB is supported in part by R21HD113234. The UCSF team is supported by the Research Allocation Program grant. JS is supported by the NIH T32 fellowship (PCORT UroGynCan T32CA251072). EJP receives funding from R01HG010863, R01AI152183, U01AI154659, the NHMRC of Australia, SJS Research Fund and the Angela Anderson Research fund. ZQ is supported by the NIH NIDDK DiabDocs K12DK133995 and a Larry L Hillblom Foundation Start Up Grant. The Vanderbilt University Medical Center dataset used in this study is supported by numerous funding sources including UL1TR002243, UL1TR000445, and UL1RR024975. The analysis of Roche data included completed clinical trials funded by Roche.

## Disclosures

DBJ has served on advisory boards or as a consultant for AstraZeneca, BMS, The Jackson Laboratory, Merck, Mosaic ImmunoEngineering, Novartis, Pfizer, and Teiko, and has received research funding from BMS and Incyte, and has patents pending for use of MHC-II as a biomarker for immune checkpoint inhibitor response, and abatacept as treatment for immune-related adverse events. EJP has served as a consultant for Janssen, Rapt, Servier, Espirion, Verve, Elion and UpToDate outside of the submitted work, and receives royalties from UpToDate. GSC and RM are employees and shareholders of Roche. They received support for preparation of this manuscript, and are co-inventors on patents filed by Roche related to use of atezolizumab. ZQ has consulted for Novartis and Sanofi.

## Acknowledgements

We thank the staff of the Vanderbilt’s Institute for Clinical and Translational Research (VICTR) and HealthIT at Vanderbilt University Medical Center (VUMC) for facilitating access to the Synthetic Derivative and VUMC-managed generative AI tools. We thank the staff of the Bakar Computational Health Sciences Institute, UCSF Information Commons, and the Center for Data-driven Insights and Innovation within the University of California Health system, as well as the UCSF AI Tiger Team, Academic Research Services, Research Information Technology, and the Chancellor’s Task Force for Generative AI for their software development, analytical and technical support related to the use of Versa API gateway (the UCSF secure implementation of large language models and generative AI via API gateway), Versa chat (the chat user interface), and related data asset and services. We would also like to acknowledge patients, families, investigators, and site staff who performed the Roche-sponsored clinical trials that were included in this analysis. Significant contributions were made by Christian W. Thorball and He Xu, both from Precision Medicine Unit, Lausanne University Hospital and University of Lausanne, Lausanne, Switzerland.

## Notes

### Author Declarations

The institutional review boards at Vanderbilt University Medical Center and University of California San Francisco approved this study, and all internal processes were followed to enable secondary use of data from Roche sponsored studies for this analysis.

