## Supplemental Figures and Tables for "irAE-GPT: Leveraging large language models to identify immune-related adverse events in electronic health records and clinical trial datasets"

**Figure S1** Workflow overview for extracting the Roche dataset.

**Figure S2** Trends of micro-averaged precision, recall, and F1 scores achieved by GPT-3.5 for various threshold values on the VUMC dataset.

**Figure S3** Trends of micro-averaged precision, recall, and F1 scores achieved by GPT-4 for various threshold values on the VUMC dataset.

**Figure S4** Trends of micro-averaged precision, recall, and F1 scores achieved by GPT-4o for various threshold values on the VUMC dataset.

### Supplemental Tables

**Table S1** Organ level classification of irAEs across VUMC, UCSF, and Roche datasets.

**Table S2** Examples of irAE synsets used in prompt development.

**Table S3** Annotation counts of irAEs in the VUMC dataset.

**Table S4** Annotation counts of irAEs in the UCSF dataset.

**Table S5** Annotation counts of irAEs in the Roche dataset.

**Table S6** Patient-level evaluation for the identification of irAEs and their corresponding categories in the VUMC dataset.

**Table S7** Patient-level evaluation for the identification of irAEs and their corresponding categories in the UCSF dataset.

**Table S8** Patient-level evaluation for the identification of irAEs and their corresponding categories in the Roche dataset.

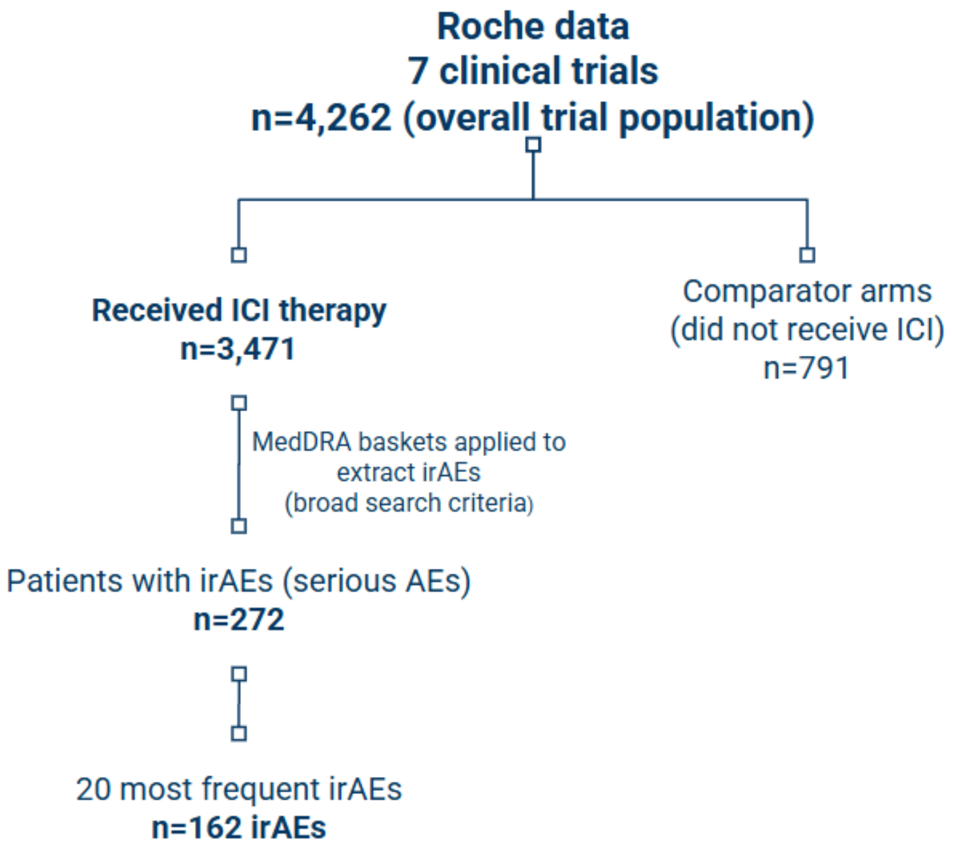

**Figure S1** Workflow overview for extracting the Roche dataset.

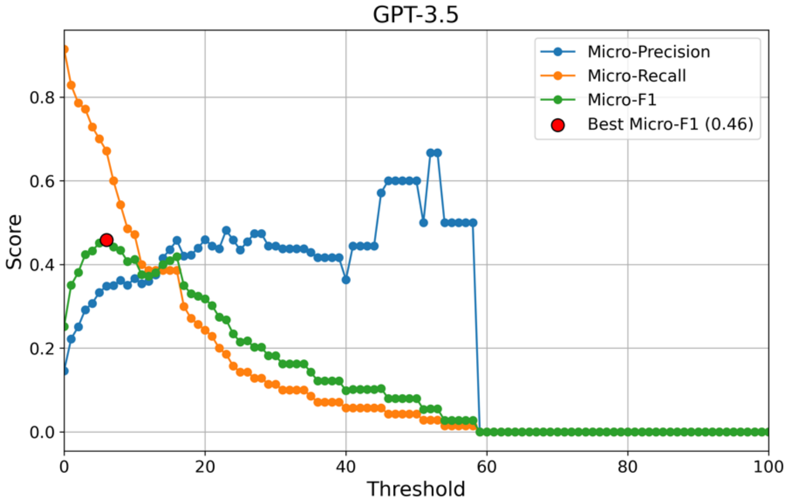

**Figure S2** Trends of micro-averaged precision, recall, and F1 scores achieved by GPT-3.5 for various threshold values on the VUMC dataset.

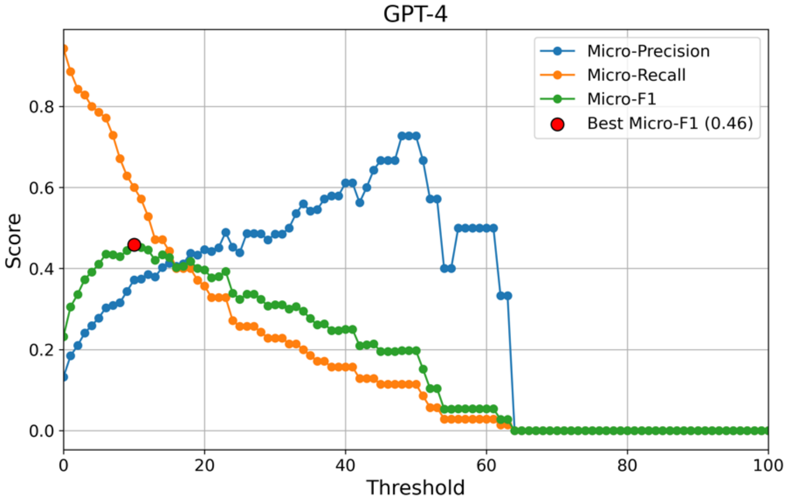

**Figure S3** Trends of micro-averaged precision, recall, and F1 scores achieved by GPT-4 for various threshold values on the VUMC dataset.

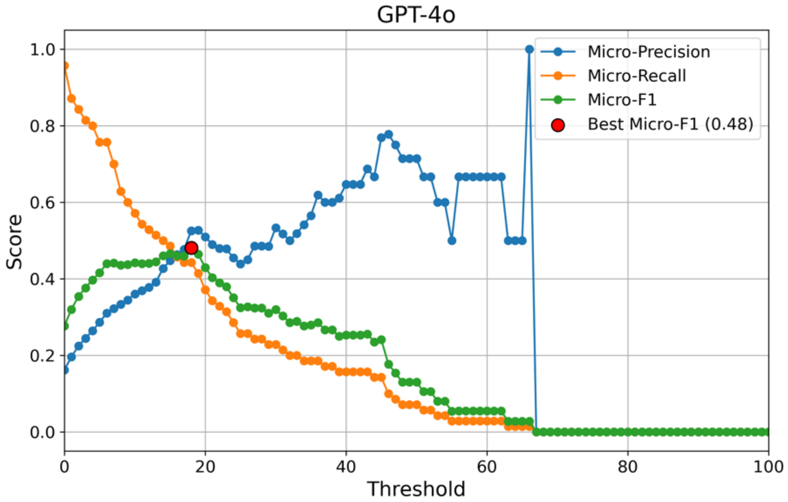

**Figure S4** Trends of micro-averaged precision, recall, and F1 scores achieved by GPT-4o for various threshold values on the VUMC dataset.

**Table S1** Organ level classification of irAEs across VUMC, UCSF, and Roche datasets.

| irAE Category | irAEs |
|---|---|
| Cardiovascular | Myocarditis, Atrial fibrillation, Pericardial effusion, Cardiac failure congestive, Pericarditis |
| Dermatologic | Rash, Dermatitis, Bullous pemphigoid, Psoriasis, Vitiligo, Itching, Flushing, Stevens-Johnson syndrome (SJS), toxic epidermal necrolysis (TEN) |
| Endocrine | Hypothyroid, Hypophysitis, Adrenal insufficiency, Diabetic ketoacidosis (DKA), Thyroiditis, Thyrotoxicosis, Hypothyroid with hyperthyroid, Hypothyroid without hyperthyroid |
| Gastrointestinal | Colitis, Hepatitis, Mucositis, Pancreatitis, Gastritis, Duodenitis, Enteritis, Ileus, Diarrhea, Stomatitis, Esophagitis |
| Hematological | Immune thrombocytopenic purpura (ITP), Neutropenia, Hemolytic anemia, Aplastic anemia, Pure red cell aplasia (PRCA), Thrombocytopenia, Hyponatraemia |
| Musculoskeletal and Rheumatologic | Arthritis, Arthralgia, Xerostomia, Rheumatoid arthritis flare (RA flare), Myositis, Lupus flare, Joint pain, Myalgia, Sarcoid, Retinal vasculitis, Ascending flaccid paralysis, Stills disease |
| Neurologic | Neuropathy, Encephalitis, Myasthenia gravis (MG), Bell's palsy, Meningitis, Uveitis, Guillain-Barre (GBS) |
| Other | Fever, Edema, Lymphadenopathy, Vaginitis, Infusion reaction, Orchitis, Pyrexia |
| Pulmonary | Pneumonitis, Wheezing, Influenza, ARDS, Pleuritis |
| Renal | Nephritis, Nephrotic syndrome, Lithiasis, Bladder tamponade, Renal failure |

**Table S2** Examples of irAE synsets used in prompt development.

| irAE | Synset |
|---|---|
| ARDS | acute respiratory distress syndrome |
| GBS | Guillain-Barre |
| Neuropathy | neurotox, neurotoxicity |
| SJS | Stevens-Johnson syndrome |
| TEN | toxic epidermal necrolysis |

**Table S3** Annotation counts of irAEs in the VUMC dataset (left column). A subset of 100 patients was used for patient-level evaluation (middle column) and a subset of 20 patients was annotated at note-level and used for note-level evaluation (right column).

| **Entire dataset with patient-level irAE annotations<br>Total patients = 747** | |
|---|---|
| **irAE** | **Patients** |
| Adrenal insufficiency | 24 |
| Aplastic anemia | 1 |
| Arthralgia | 17 |
| Arthritis | 19 |
| Bell's palsy | 1 |
| Bullous pemphigoid | 3 |
| Colitis | 107 |
| Dermatitis | 18 |
| Diarrhea | 1 |
| Diabetic ketoacidosis (DKA) | 5 |
| Duodenitis | 2 |
| Edema | 1 |
| Encephalitis | 7 |
| Enteritis | 1 |
| Fever | 4 |
| Flushing | 1 |
| Gastritis | 4 |
| Guillain-Barre | 1 |
| Hemolytic anemia | 1 |
| Hepatitis | 47 |
| Hypophysitis | 33 |
| Hypothyroid | 49 |
| Ileus | 1 |
| Infusion reaction | 1 |
| Itching | 1 |
| Immune thrombocytopenic purpura (ITP) | 3 |
| Joint pain | 1 |
| Lupus flare | 1 |
| Lymphadenopathy | 1 |
| Mengingitis | 1 |
| Mucositis | 7 |
| Myalgia | 1 |
| Myasthenia gravis (MG) | 3 |
| Myocarditis | 3 |
| Myositis | 5 |
| Nephritis | 9 |
| Nephrotic syndrome | 1 |
| Neuropathy | 10 |
| Neutropenia | 2 |
| Orchitis | 1 |
| Pancreatitis | 6 |
| Pneumonitis | 70 |
| Pure red cell aplasia (PRCA) | 1 |
| Psoriasis | 3 |
| Rheumatoid arthritis flare (RA flare) | 5 |
| Rash | 102 |
| Retinal vasculitis | 1 |
| Sarcoid | 1 |
| Stomatitis | 1 |
| Thrombocytopenia | 1 |
| Thyroiditis | 3 |
| Thyrotoxicosis | 1 |
| Uveitis | 1 |
| Vaginitis | 1 |
| Vitiligo | 1 |
| Wheezing | 1 |
| Xerostomia | 5 |
| **None*** | **292** |

| **Dataset for patient-level evaluation<br>Total patients = 100** | |
|---|---|
| **irAE** | **Patients** |
| Adrenal insufficiency | 5 |
| Arthritis | 6 |
| Bullous pemphigoid | 1 |
| Colitis | 9 |
| Dermatitis | 3 |
| Diabetic ketoacidosis (DKA) | 1 |
| Diarrhea | 1 |
| Hemolytic anemia | 1 |
| Hepatitis | 3 |
| Hypophysitis | 2 |
| Hypothyroid | 6 |
| Lupus flare | 1 |
| Mucositis | 3 |
| Myocarditis | 1 |
| Myositis | 2 |
| Nephritis | 2 |
| Neuropathy | 1 |
| Pancreatitis | 1 |
| Pneumonitis | 8 |
| Psoriasis | 1 |
| Rash | 10 |
| Rheumatoid arthritis flare (RA flare) | 1 |
| Thyrotoxicosis | 1 |
| **None*** | **42** |

| **Dataset for note-level evaluation<br>Total notes = 667** | |
|---|---|
| **irAE** | **Notes** |
| Adrenal insufficiency | 21 |
| Arthralgia | 21 |
| Arthritis | 7 |
| Colitis | 26 |
| Duodenitis | 15 |
| Fever | 2 |
| Hepatitis | 21 |
| Hypophysitis | 12 |
| Hypothyroid | 44 |
| Joint pain | 3 |
| Mucositis | 11 |
| Myalgia | 2 |
| Myasthenia gravis (MG) | 46 |
| Neuropathy | 49 |
| Pancreatitis | 13 |
| Pneumonitis | 6 |
| Rash | 27 |
| **None*** | **407** |

* 'None' represents patients not experiencing any irAE or patient notes not describing any specific irAE experienced by the patient. Notably, all the patients in the VUMC dataset underwent ICI therapy and may have experienced none, one, or multiple irAEs.

**Table S4** Annotation counts of irAEs in the UCSF dataset.

| Dataset for patient-level evaluation<br>Total patients = 70 | |
|---|---|
| **irAE** | **Patients** |
| ARDS | 1 |
| Arthritis | 3 |
| Colitis | 23 |
| DM | 5 |
| Encephalitis | 2 |
| Enteritis | 2 |
| Esophagitis | 1 |
| Fever | 4 |
| GBS | 1 |
| Hepatitis | 15 |
| Hypophysitis | 4 |
| Hypothyroid with hyperthyroid | 5 |
| Hypothyroid without hyperthyroid | 3 |
| Mucositis | 2 |
| Myasthenia gravis | 2 |
| Myocarditis | 2 |
| Myositis | 3 |
| Pancreatitis | 1 |
| Pericardial effusion | 1 |
| Pericarditis | 1 |
| Pleuritis | 2 |
| Pneumonitis | 15 |
| Rash | 7 |
| SJS | 1 |
| Stills Disease | 1 |
| TEN | 1 |

**Table S5** Annotation counts of irAEs in the Roche dataset.

| Dataset for patient-level evaluation<br>Total patients = 272 | |
|---|---|
| **irAE** | **Patients** |
| Adrenal insufficiency | 10 |
| Ascending flaccid paralysis | 1 |
| Atrial fibrillation | 1 |
| Bladder tamponade* | 1 |
| Cardiac failure congestive | 1 |
| Colitis | 24 |
| Enteritis | 1 |
| Hepatitis | 10 |
| Hyperthyroidism | 4 |
| Hyponatraemia | 4 |
| Hypothyroidism | 8 |
| Influenza* | 3 |
| Lithiasis | 1 |
| Meningitis | 7 |
| Pancreatitis | 8 |
| Pericardial effusion | 9 |
| Pneumonitis | 41 |
| Pyrexia | 11 |
| Rash | 15 |
| Renal failure | 2 |

* Certain adverse events may not qualify as irAEs but might instead be indirectly associated with ICI therapy.

**Table S6** Patient-level evaluation for the identification of irAEs and their corresponding categories in the VUMC dataset.

| | GPT-3.5 | | | | GPT-4 | | | | GPT-4o | | | |
|---|---|---|---|---|---|---|---|---|---|---|---|---|
| **irAE** | **P** | **R** | **S** | **F1** | **P** | **R** | **S** | **F1** | **P** | **R** | **S** | **F1** |
| Adrenal insufficiency | 0.625 | 1.000 | 0.968 | 0.769 | 0.667 | 0.800 | 0.979 | 0.727 | 0.667 | 0.400 | 0.989 | 0.500 |
| Arthritis | 0.333 | 0.167 | 0.979 | 0.222 | 0.333 | 0.167 | 0.979 | 0.222 | 0.000 | 0.000 | 1.000 | 0.000 |
| Bullous pemphigoid | 1.000 | 1.000 | 1.000 | 1.000 | 1.000 | 1.000 | 1.000 | 1.000 | 1.000 | 1.000 | 1.000 | 1.000 |
| Colitis | 0.667 | 0.444 | 0.978 | 0.533 | 0.500 | 0.111 | 0.989 | 0.182 | 1.000 | 0.111 | 1.000 | 0.200 |
| Dermatitis | 0.000 | 0.000 | 0.990 | 0.000 | 0.000 | 0.000 | 1.000 | 0.000 | 0.000 | 0.000 | 1.000 | 0.000 |
| Diabetic ketoacidosis | 0.500 | 1.000 | 0.990 | 0.667 | 1.000 | 1.000 | 1.000 | 1.000 | 1.000 | 1.000 | 1.000 | 1.000 |
| Diarrhea | 0.048 | 1.000 | 0.798 | 0.091 | 0.056 | 1.000 | 0.828 | 0.105 | 0.167 | 1.000 | 0.949 | 0.286 |
| Hemolytic anemia | 1.000 | 1.000 | 1.000 | 1.000 | 1.000 | 1.000 | 1.000 | 1.000 | 1.000 | 1.000 | 1.000 | 1.000 |
| Hepatitis | 1.000 | 0.667 | 1.000 | 0.800 | 0.500 | 0.333 | 0.990 | 0.400 | 1.000 | 0.333 | 1.000 | 0.500 |
| Hypophysitis | 0.667 | 1.000 | 0.990 | 0.800 | 0.667 | 1.000 | 0.990 | 0.800 | 0.500 | 0.500 | 0.990 | 0.500 |
| Hypothyroid | 0.263 | 0.833 | 0.851 | 0.400 | 0.250 | 0.500 | 0.904 | 0.333 | 0.333 | 0.500 | 0.936 | 0.400 |
| Lupus flare | 1.000 | 1.000 | 1.000 | 1.000 | 1.000 | 1.000 | 1.000 | 1.000 | 1.000 | 1.000 | 1.000 | 1.000 |
| Mucositis | 0.000 | 0.000 | 1.000 | 0.000 | 1.000 | 0.667 | 1.000 | 0.800 | 1.000 | 0.333 | 1.000 | 0.500 |
| Myocarditis | 0.500 | 1.000 | 0.990 | 0.667 | 0.500 | 1.000 | 0.990 | 0.667 | 0.500 | 1.000 | 0.990 | 0.667 |
| Myositis | 0.500 | 0.500 | 0.990 | 0.500 | 0.500 | 0.500 | 0.990 | 0.500 | 0.000 | 0.000 | 1.000 | 0.000 |
| Nephritis | 1.000 | 0.500 | 1.000 | 0.667 | 1.000 | 0.500 | 1.000 | 0.667 | 1.000 | 0.500 | 1.000 | 0.667 |
| Neuropathy | 0.250 | 1.000 | 0.970 | 0.400 | 0.500 | 1.000 | 0.990 | 0.667 | 0.500 | 1.000 | 0.990 | 0.667 |
| Pancreatitis | 0.000 | 0.000 | 0.990 | 0.000 | 0.000 | 0.000 | 1.000 | 0.000 | 0.000 | 0.000 | 1.000 | 0.000 |
| Pneumonitis | 0.800 | 1.000 | 0.978 | 0.889 | 0.800 | 1.000 | 0.978 | 0.889 | 0.857 | 0.750 | 0.989 | 0.800 |
| Psoriasis | 1.000 | 1.000 | 1.000 | 1.000 | 0.500 | 1.000 | 0.990 | 0.667 | 0.500 | 1.000 | 0.990 | 0.667 |
| Rash | 0.190 | 0.800 | 0.622 | 0.308 | 0.200 | 0.800 | 0.644 | 0.320 | 0.353 | 0.600 | 0.878 | 0.444 |
| Rheumatoid arthritis flare | 0.500 | 1.000 | 0.990 | 0.667 | 1.000 | 1.000 | 1.000 | 1.000 | 0.000 | 0.000 | 1.000 | 0.000 |
| Thyrotoxicosis | 0.500 | 1.000 | 0.990 | 0.667 | 1.000 | 1.000 | 1.000 | 1.000 | 1.000 | 1.000 | 1.000 | 1.000 |
| **micro avg** | 0.348 | 0.671 | 0.961 | 0.459 | 0.372 | 0.600 | 0.968 | 0.459 | 0.525 | 0.443 | 0.987 | 0.481 |
| **macro avg** | 0.537 | 0.735 | 0.959 | 0.567 | 0.607 | 0.712 | 0.967 | 0.606 | 0.582 | 0.566 | 0.987 | 0.513 |
| **irAE Category** | | | | | | | | | | | | |
| Cardiovascular | 0.500 | 1.000 | 0.990 | 0.667 | 0.500 | 1.000 | 0.990 | 0.667 | 0.500 | 1.000 | 0.990 | 0.667 |
| Dermatologic | 0.302 | 0.867 | 0.647 | 0.448 | 0.317 | 0.867 | 0.671 | 0.464 | 0.500 | 0.667 | 0.882 | 0.571 |
| Endocrine | 0.519 | 0.933 | 0.847 | 0.667 | 0.579 | 0.733 | 0.906 | 0.647 | 0.615 | 0.533 | 0.941 | 0.571 |
| Gastrointestinal | 0.480 | 0.706 | 0.843 | 0.571 | 0.429 | 0.529 | 0.855 | 0.474 | 0.667 | 0.353 | 0.964 | 0.462 |
| Hematological | 1.000 | 1.000 | 1.000 | 1.000 | 1.000 | 1.000 | 1.000 | 1.000 | 1.000 | 1.000 | 1.000 | 1.000 |
| Musculoskeletal and Rheumatologic | 0.714 | 0.500 | 0.978 | 0.588 | 0.571 | 0.400 | 0.967 | 0.471 | 1.000 | 0.100 | 1.000 | 0.182 |
| Neurologic | 0.250 | 1.000 | 0.970 | 0.400 | 0.500 | 1.000 | 0.990 | 0.667 | 0.500 | 1.000 | 0.990 | 0.667 |
| Pulmonary | 0.800 | 1.000 | 0.978 | 0.889 | 0.800 | 1.000 | 0.978 | 0.889 | 0.857 | 0.750 | 0.989 | 0.800 |
| Renal | 1.000 | 0.500 | 1.000 | 0.667 | 1.000 | 0.500 | 1.000 | 0.667 | 1.000 | 0.500 | 1.000 | 0.667 |
| **micro avg** | 0.467 | 0.800 | 0.923 | 0.589 | 0.471 | 0.700 | 0.934 | 0.563 | 0.625 | 0.500 | 0.975 | 0.556 |
| **macro avg** | 0.618 | 0.834 | 0.917 | 0.655 | 0.633 | 0.781 | 0.929 | 0.661 | 0.738 | 0.656 | 0.973 | 0.621 |

P, precision (positive predictive value); R, recall (sensitivity); S, specificity; F1, F1-measure.

**Table S7** Patient-level evaluation for the identification of irAEs and their corresponding categories in the UCSF dataset.

| | GPT-3.5 | | | | GPT-4 | | | | GPT-4o | | | |
|---|---|---|---|---|---|---|---|---|---|---|---|---|
| **irAE** | **P** | **R** | **S** | **F1** | **P** | **R** | **S** | **F1** | **P** | **R** | **S** | **F1** |
| Rash | 0.400 | 0.857 | 0.857 | 0.545 | 0.273 | 0.857 | 0.746 | 0.414 | 0.300 | 0.857 | 0.778 | 0.444 |
| SJS | 1.000 | 1.000 | 1.000 | 1.000 | 1.000 | 1.000 | 1.000 | 1.000 | 1.000 | 1.000 | 1.000 | 1.000 |
| TEN | 0.000 | 0.000 | 1.000 | 0.000 | 0.000 | 0.000 | 1.000 | 0.000 | 0.000 | 0.000 | 1.000 | 0.000 |
| Stills Disease | 0.500 | 1.000 | 0.986 | 0.667 | 0.500 | 1.000 | 0.986 | 0.667 | 0.500 | 1.000 | 0.986 | 0.667 |
| Colitis | 0.688 | 0.957 | 0.787 | 0.800 | 0.564 | 0.957 | 0.638 | 0.710 | 0.618 | 0.913 | 0.723 | 0.737 |
| Hepatitis | 0.800 | 0.533 | 0.964 | 0.640 | 0.647 | 0.733 | 0.891 | 0.688 | 0.733 | 0.733 | 0.927 | 0.733 |
| Pancreatitis | 0.500 | 1.000 | 0.986 | 0.667 | 0.250 | 1.000 | 0.957 | 0.400 | 1.000 | 1.000 | 1.000 | 1.000 |
| Mucositis | 0.500 | 1.000 | 0.971 | 0.667 | 0.333 | 1.000 | 0.941 | 0.500 | 0.333 | 1.000 | 0.941 | 0.500 |
| Esophagitis | 0.500 | 1.000 | 0.986 | 0.667 | 0.500 | 1.000 | 0.986 | 0.667 | 0.333 | 1.000 | 0.971 | 0.500 |
| Enteritis | 0.286 | 1.000 | 0.926 | 0.444 | 0.250 | 1.000 | 0.912 | 0.400 | 0.250 | 1.000 | 0.912 | 0.400 |
| Pneumonitis | 0.706 | 0.800 | 0.909 | 0.750 | 0.650 | 0.867 | 0.873 | 0.743 | 0.722 | 0.867 | 0.909 | 0.788 |
| ARDS | 0.000 | 0.000 | 0.928 | 0.000 | 0.000 | 0.000 | 0.986 | 0.000 | 0.000 | 0.000 | 0.986 | 0.000 |
| Pleuritis | 0.000 | 0.000 | 1.000 | 0.000 | 0.000 | 0.000 | 1.000 | 0.000 | 0.000 | 0.000 | 1.000 | 0.000 |
| Hypothyroid with hyperthyroid | 0.235 | 0.800 | 0.800 | 0.364 | 0.208 | 1.000 | 0.708 | 0.345 | 0.313 | 1.000 | 0.831 | 0.476 |
| Hypothyroid without hyperthyroid | 0.214 | 1.000 | 0.836 | 0.353 | 0.130 | 1.000 | 0.701 | 0.231 | 0.200 | 1.000 | 0.821 | 0.333 |
| Hypophysitis | 0.400 | 1.000 | 0.909 | 0.571 | 0.364 | 1.000 | 0.894 | 0.533 | 0.500 | 1.000 | 0.939 | 0.667 |
| DM | 0.286 | 0.800 | 0.846 | 0.421 | 0.278 | 1.000 | 0.800 | 0.435 | 0.357 | 1.000 | 0.862 | 0.526 |
| Arthritis | 0.167 | 0.667 | 0.851 | 0.267 | 0.167 | 1.000 | 0.776 | 0.286 | 0.250 | 0.667 | 0.910 | 0.364 |
| Myositis | 1.000 | 1.000 | 1.000 | 1.000 | 0.750 | 1.000 | 0.985 | 0.857 | 0.750 | 1.000 | 0.985 | 0.857 |
| Myocarditis | 0.667 | 1.000 | 0.985 | 0.800 | 0.500 | 1.000 | 0.971 | 0.667 | 0.500 | 1.000 | 0.971 | 0.667 |
| Pericarditis | 0.333 | 1.000 | 0.971 | 0.500 | 0.333 | 1.000 | 0.971 | 0.500 | 0.500 | 1.000 | 0.986 | 0.667 |
| Pericardial effusion | 0.250 | 1.000 | 0.957 | 0.400 | 0.100 | 1.000 | 0.870 | 0.182 | 0.333 | 1.000 | 0.971 | 0.500 |
| GBS | 1.000 | 1.000 | 1.000 | 1.000 | 1.000 | 1.000 | 1.000 | 1.000 | 1.000 | 1.000 | 1.000 | 1.000 |
| Myasthenia gravis | 0.667 | 1.000 | 0.985 | 0.800 | 1.000 | 1.000 | 1.000 | 1.000 | 1.000 | 1.000 | 1.000 | 1.000 |
| Fever | 0.235 | 1.000 | 0.803 | 0.381 | 0.148 | 1.000 | 0.652 | 0.258 | 0.190 | 1.000 | 0.742 | 0.320 |
| Encephalitis | 1.000 | 0.500 | 1.000 | 0.667 | 0.667 | 1.000 | 0.985 | 0.800 | 0.500 | 0.500 | 0.985 | 0.500 |
| **micro avg** | 0.442 | 0.815 | 0.935 | 0.573 | 0.356 | 0.889 | 0.898 | 0.508 | 0.445 | 0.861 | 0.932 | 0.587 |
| **macro avg** | 0.474 | 0.804 | 0.932 | 0.553 | 0.408 | 0.862 | 0.893 | 0.511 | 0.469 | 0.828 | 0.928 | 0.563 |
| **irAE Category** | | | | | | | | | | | | |
| Dermatologic | 0.233 | 0.875 | 0.629 | 0.368 | 0.280 | 0.875 | 0.710 | 0.424 | 0.292 | 0.875 | 0.726 | 0.438 |
| Gastrointestinal | 0.786 | 0.917 | 0.735 | 0.846 | 0.700 | 0.972 | 0.559 | 0.814 | 0.773 | 0.944 | 0.706 | 0.850 |
| Pulmonary | 0.706 | 0.706 | 0.906 | 0.706 | 0.667 | 0.824 | 0.868 | 0.737 | 0.737 | 0.824 | 0.906 | 0.778 |
| Endocrine | 0.438 | 0.933 | 0.673 | 0.596 | 0.385 | 1.000 | 0.564 | 0.556 | 0.484 | 1.000 | 0.709 | 0.652 |
| Musculoskeletal and Rheumatologic | 0.353 | 0.857 | 0.825 | 0.500 | 0.304 | 1.000 | 0.746 | 0.467 | 0.429 | 0.857 | 0.873 | 0.571 |
| Cardiovascular | 0.500 | 1.000 | 0.939 | 0.667 | 0.222 | 1.000 | 0.788 | 0.364 | 0.500 | 1.000 | 0.939 | 0.667 |
| Neurologic | 0.400 | 0.800 | 0.908 | 0.533 | 0.357 | 1.000 | 0.862 | 0.526 | 0.444 | 0.800 | 0.923 | 0.571 |
| Other | 0.235 | 1.000 | 0.803 | 0.381 | 0.148 | 1.000 | 0.652 | 0.258 | 0.190 | 1.000 | 0.742 | 0.320 |
| **micro avg** | 0.486 | 0.875 | 0.808 | 0.625 | 0.419 | 0.948 | 0.728 | 0.581 | 0.518 | 0.917 | 0.823 | 0.662 |
| **macro avg** | 0.456 | 0.886 | 0.802 | 0.575 | 0.383 | 0.959 | 0.718 | 0.518 | 0.481 | 0.913 | 0.816 | 0.606 |

P, precision (positive predictive value); R, recall (sensitivity); S, specificity; F1, F1-measure.

**Table S8** Patient-level evaluation for the identification of irAEs and their corresponding categories in the Roche dataset.

| | GPT-3.5 | | | | GPT-4 | | | | GPT-4o | | | |
|---|---|---|---|---|---|---|---|---|---|---|---|---|
| **irAE** | **P** | **R** | **S** | **F1** | **P** | **R** | **S** | **F1** | **P** | **R** | **S** | **F1** |
| Colitis | 0.571 | 0.833 | 0.940 | 0.678 | 0.545 | 1.000 | 0.919 | 0.706 | 0.605 | 0.958 | 0.940 | 0.742 |
| Pneumonitis | 0.712 | 0.902 | 0.935 | 0.796 | 0.719 | 1.000 | 0.931 | 0.837 | 0.774 | 1.000 | 0.948 | 0.872 |
| Hepatitis | 0.333 | 1.000 | 0.924 | 0.500 | 0.208 | 1.000 | 0.855 | 0.345 | 0.250 | 1.000 | 0.885 | 0.400 |
| Adrenal insufficiency | 0.643 | 0.900 | 0.981 | 0.750 | 0.667 | 1.000 | 0.981 | 0.800 | 0.769 | 1.000 | 0.989 | 0.870 |
| Pyrexia | 0.188 | 0.545 | 0.900 | 0.279 | 0.196 | 0.909 | 0.843 | 0.323 | 0.196 | 1.000 | 0.828 | 0.328 |
| Atrial fibrillation | 0.077 | 1.000 | 0.956 | 0.143 | 0.071 | 1.000 | 0.952 | 0.133 | 0.143 | 1.000 | 0.978 | 0.250 |
| Influenza | 0.500 | 1.000 | 0.989 | 0.667 | 0.750 | 1.000 | 0.996 | 0.857 | 0.000 | 0.000 | 1.000 | 0.000 |
| Rash | 0.400 | 0.800 | 0.930 | 0.533 | 0.280 | 0.933 | 0.860 | 0.431 | 0.438 | 0.933 | 0.930 | 0.596 |
| Lithiasis | 0.125 | 1.000 | 0.974 | 0.222 | 0.500 | 1.000 | 0.996 | 0.667 | 0.000 | 0.000 | 1.000 | 0.000 |
| Pericardial effusion | 0.500 | 1.000 | 0.966 | 0.667 | 0.450 | 1.000 | 0.958 | 0.621 | 0.538 | 0.778 | 0.977 | 0.636 |
| Ascending flaccid paralysis | 0.500 | 1.000 | 0.996 | 0.667 | 0.500 | 1.000 | 0.996 | 0.667 | 1.000 | 1.000 | 1.000 | 1.000 |
| Hypothyroidism | 0.222 | 1.000 | 0.894 | 0.364 | 0.182 | 1.000 | 0.864 | 0.308 | 0.333 | 1.000 | 0.939 | 0.500 |
| Hyperthyroidism | 0.800 | 1.000 | 0.996 | 0.889 | 0.500 | 1.000 | 0.985 | 0.667 | 0.500 | 1.000 | 0.985 | 0.667 |
| Cardiac failure congestive | 0.000 | 0.000 | 0.993 | 0.000 | 0.500 | 1.000 | 0.996 | 0.667 | 0.500 | 1.000 | 0.996 | 0.667 |
| Hyponatraemia | 0.250 | 0.500 | 0.978 | 0.333 | 0.190 | 1.000 | 0.937 | 0.320 | 0.222 | 1.000 | 0.948 | 0.364 |
| Bladder tamponade | 0.000 | 0.000 | 1.000 | 0.000 | 1.000 | 1.000 | 1.000 | 1.000 | 1.000 | 1.000 | 1.000 | 1.000 |
| Meningitis | 0.714 | 0.714 | 0.992 | 0.714 | 0.875 | 1.000 | 0.996 | 0.933 | 0.875 | 1.000 | 0.996 | 0.933 |
| Renal failure | 0.143 | 0.500 | 0.978 | 0.222 | 0.056 | 1.000 | 0.874 | 0.105 | 0.182 | 1.000 | 0.967 | 0.308 |
| Enteritis | 0.000 | 0.000 | 0.982 | 0.000 | 0.111 | 1.000 | 0.970 | 0.200 | 0.000 | 0.000 | 0.993 | 0.000 |
| Pancreatitis | 0.889 | 1.000 | 0.996 | 0.941 | 0.889 | 1.000 | 0.996 | 0.941 | 1.000 | 0.875 | 1.000 | 0.933 |
| **micro avg** | 0.429 | 0.846 | 0.966 | 0.570 | 0.360 | 0.988 | 0.946 | 0.527 | 0.455 | 0.938 | 0.966 | 0.613 |
| **macro avg** | 0.378 | 0.735 | 0.965 | 0.468 | 0.460 | 0.992 | 0.945 | 0.576 | 0.466 | 0.827 | 0.965 | 0.553 |
| **irAE Category** | | | | | | | | | | | | |
| Cardiovascular | 0.323 | 0.909 | 0.920 | 0.476 | 0.324 | 1.000 | 0.912 | 0.489 | 0.409 | 0.818 | 0.950 | 0.545 |
| Dermatologic | 0.400 | 0.800 | 0.930 | 0.533 | 0.280 | 0.933 | 0.860 | 0.431 | 0.438 | 0.933 | 0.930 | 0.596 |
| Endocrine | 0.408 | 0.952 | 0.884 | 0.571 | 0.356 | 1.000 | 0.849 | 0.525 | 0.525 | 1.000 | 0.924 | 0.689 |
| Gastrointestinal | 0.507 | 0.905 | 0.839 | 0.650 | 0.396 | 1.000 | 0.722 | 0.568 | 0.471 | 0.952 | 0.804 | 0.630 |
| Hematological | 0.250 | 0.500 | 0.978 | 0.333 | 0.190 | 1.000 | 0.937 | 0.320 | 0.222 | 1.000 | 0.948 | 0.364 |
| Musculoskeletal and Rheumatologic | 0.500 | 1.000 | 0.996 | 0.667 | 0.500 | 1.000 | 0.996 | 0.667 | 1.000 | 1.000 | 1.000 | 1.000 |
| Neurologic | 0.714 | 0.714 | 0.992 | 0.714 | 0.875 | 1.000 | 0.996 | 0.933 | 0.875 | 1.000 | 0.996 | 0.933 |
| Other | 0.188 | 0.545 | 0.900 | 0.279 | 0.196 | 0.909 | 0.843 | 0.323 | 0.196 | 1.000 | 0.828 | 0.328 |
| Pulmonary | 0.702 | 0.909 | 0.925 | 0.792 | 0.721 | 1.000 | 0.925 | 0.838 | 0.774 | 0.932 | 0.947 | 0.845 |
| Renal | 0.143 | 0.500 | 0.955 | 0.222 | 0.103 | 1.000 | 0.869 | 0.186 | 0.250 | 0.750 | 0.966 | 0.375 |
| **micro avg** | 0.446 | 0.850 | 0.934 | 0.585 | 0.367 | 0.988 | 0.893 | 0.535 | 0.462 | 0.944 | 0.931 | 0.620 |
| **macro avg** | 0.413 | 0.774 | 0.932 | 0.524 | 0.394 | 0.984 | 0.891 | 0.528 | 0.516 | 0.939 | 0.929 | 0.631 |

P, precision (positive predictive value); R, recall (sensitivity); S, specificity; F1, F1-measure.

**Table S9** Clinical note excerpts highlighting cases where LLMs misclassified adverse events as irAEs.

| Adverse event misclassified as irAE | Note excerpt | Reason for misclassification |
|---|---|---|
| Hypothyroidism | *Past medical history: hypothyroidism* | Event happened in the past due to unknown cause |
| Rash | *side effects of chemotherapy including but not limited to alopecia, fatigue, nausea and vomiting, weight loss, change in taste, rash* | The adverse event was caused by chemotherapy, not ICI exposure |
| Pneumonitis | *temsirolimus [DATE**] – [DATE**], discontinued due to pneumonitis* | The adverse event was caused by temsirolimus, not ICI exposure |
| Hypothyroidism | *I suspect he is trending toward hypothyroid but actually has a slightly elevated free T4 at this time* | Assertion status of the adverse event: uncertain/possible |
| Colitis | *it is difficult to determine if this is a possible pembrolizumab associated colitis* | Assertion status of the adverse event: uncertain/possible |
| Adrenal insufficiency | *will do ACTH stimulation test today as adrenal insufficiency could occur with pembrolizumab* | Assertion status of the adverse event: uncertain/possible |
| Hypothyroidism | *Family medical history: - sister: hyperthyroidism* | Assertion status of the adverse event: not associated with the patient |
